# Effectiveness of using manual pill organizers and pill reminder apps in improving medication adherence and health outcomes in the Indian elderly population receiving multiple medications (PORA-MEDAdhere): Protocol for a 2×2 factorial randomized controlled trial

**DOI:** 10.1101/2025.07.03.25330787

**Authors:** Aditi Apte, Farah Naaz Fatima, Bhupendra Solanke, Sumithra Selvam, Dhiraj Agarwal, Pooja Shridhar, Harpreet Singh, Radhika Nimkar, Rakesh Patil, Jerin Jose Cherian, Sudipto Roy

**Author notes:** **Corresponding author:** Dr Aditi Apte, KEM Hospital Research Centre, Pune-411011 **Email:**.

## Abstract

**Introduction:** Poor medication adherence is associated with poor clinical outcomes, an increase in hospitalisations, and increased mortality. This is a multi-center randomised study that evaluates the effectiveness of using a manual pill organiser (MPO) and a custom-developed pill reminder app (PRA) on medication adherence, morbidity, as well as health economic outcomes amongst Indian elderly individuals taking multiple medications.

**Objectives:** The primary objective of this study is to evaluate the impact of MPO and PRA alone or in combination in improving medication adherence amongst elderly individuals on multiple medications. The secondary objectives include the impact of interventions on the morbidity profile and health-related quality of life. The study also plans to assess the cost-effectiveness and cost-utility of improving medication adherence.

**Methods and analysis:** This is a community-based, open-label, factorial-design randomized controlled trial to be conducted across rural and urban populations at two geographically distinct sites in India. The study will enrol 752 elderly individuals aged 60-80 years, receiving three or more medications for at least six months and having access the smartphones. The participants will be randomised to receive one of the following interventions for 12 months: Control group, PRA, MPO, and MPO + PRA. All study groups would receive patient education about the importance of medication adherence. The study outcomes include the proportion of improvement in medication adherence [using medication adherence rating system (MARS-5), seven-day point prevalence of medication non-adherence and pill count]; morbidity profile; health care utilisation; health-related quality of life; cost-effectiveness and cost-utility outcomes.

**Ethics and dissemination:** The study protocol has been approved by institutional ethics committees at all three institutes. The study results for primary and secondary outcomes will be published in peer-reviewed journals.

**Trial registration number:** CTRI/2024/01/061975 [Registered on-29/01/2024].

*Strengths and limitations:*

- The study provides the first clinical evidence of the effectiveness of locally developed pill reminder apps and manual pill organisers in improving adherence in Indian elderly individuals with non-communicable diseases.
- The study incorporates rural and urban populations from two geographically different sites in India, enhancing its representativeness
- The study evaluates the intervention’s impact on the morbidity profile, healthcare utilisation, quality of life, and includes cost-effectiveness and cost-utility analyses
- The factorial design enables evaluation of the individual and combined intervention effects against the control
- Limited literacy among rural elderly people and lowlow Android phones availability are expected challenges and limitations in implementation
- The study relies on multiple outcomes to assess medication adherence due to the lack of a gold standard and the subjectivity of its measurement.

## Introduction

Noncommunicable diseases are the leading causes of death globally, disproportionately affecting low- and middle-income countries. In 2017, non-communicable diseases accounted for 4.7 million deaths and 226.8 million disability-adjusted life years (1). Medication non-adherence is extremely common globally amongst elderly individuals taking medications for chronic diseases such as diabetes and hypertension, affecting as many as 60-75% of these individuals (2,3).

Insufficient adherence to medications is associated with poor clinical outcomes, an increase in hospitalisations, and increased mortality (4). A meta-analysis shows that medication non-adherence is significantly associated with an increase in all-cause mortality, and good adherence is associated with a 21% reduced risk of long-term mortality in adults aged more than 55 years (5). Poor medication adherence is reported to be associated with 200000 premature deaths every year in Europe, thus resulting in a high health and economic cost to society (6). These numbers are likely to be higher in India, where a large proportion of people suffer from illiteracy and have limited access to health services. Further, medication non-adherence is associated with reduced health-related quality of life, especially among elderly people and people suffering from neurological diseases (7,8).

Medication non-adherence could be due to medication-related factors such as an increase in the number of medications, adverse effects, poor communication skills of the prescriber, non-affordability, and lack of medication review by the prescriber in the recent past. It can also result from several personal factors such as forgetfulness, low health literacy, lack of motivation or awareness, and problems related to vision and cognition (9)(10). These factors can be generally categorised as intentional and non-intentional. Non-intentional non-adherence is an unplanned behaviour due to factors beyond the patient’s control, and intentional non-adherence is driven by patients’ beliefs and knowledge about the disease and the medications (11).

## Rationale

Interventions such as patient education, simplification of medication regimen, and medication-taking reminders have been shown to improve medication adherence for chronic diseases (12). The intervention of patient education, especially in-person counselling, has been shown to improve medication adherence, especially due to intentional reasons (13). Technologies can provide solutions for automated reminders, and medication adherence monitoring can provide an opportunity for continuous tracking of individual medication adherence behaviour and thus help to improve non-intentional adherence. Manual pill organiser (MPO) and pill reminder mobile app (PRA) are simple and easy-to-use interventions that can be used to reduce medication non-adherence in elderly individuals (14)(15). Considering the multitude of factors involved in medication non-adherence, complex interventions including one or more of the above interventions would be useful. There is limited systematic evidence on the impact of these interventions on health outcomes and their cost-effectiveness. Also, most of the mobile applications available are not suited for the Indian context and are not available in regional languages.

This is a multi-centre randomised study that evaluates the effectiveness of using MPO and an custom-developed PRA in improving medication adherence amongst Indian elderly patients taking multiple medications. The study interventions have been pilot-tested amongst elderly individuals and have shown good feasibility and acceptability for long-term use (16).

## Objectives

The primary objective of this study will be to evaluate the impact of MPO and PRA in improving medication adherence amongst elderly patients aged 60-80 years on multiple medications. Additionally, the study plans to evaluate the impact of the use of MPO and PRA in improving morbidities, healthcare utilisation and health-related quality of life. The study also plans to evaluate the cost-effectiveness of the interventions and their impact on cost-utility outcomes. The PICOT format is shown in Figure 1.

**Figure 1:**
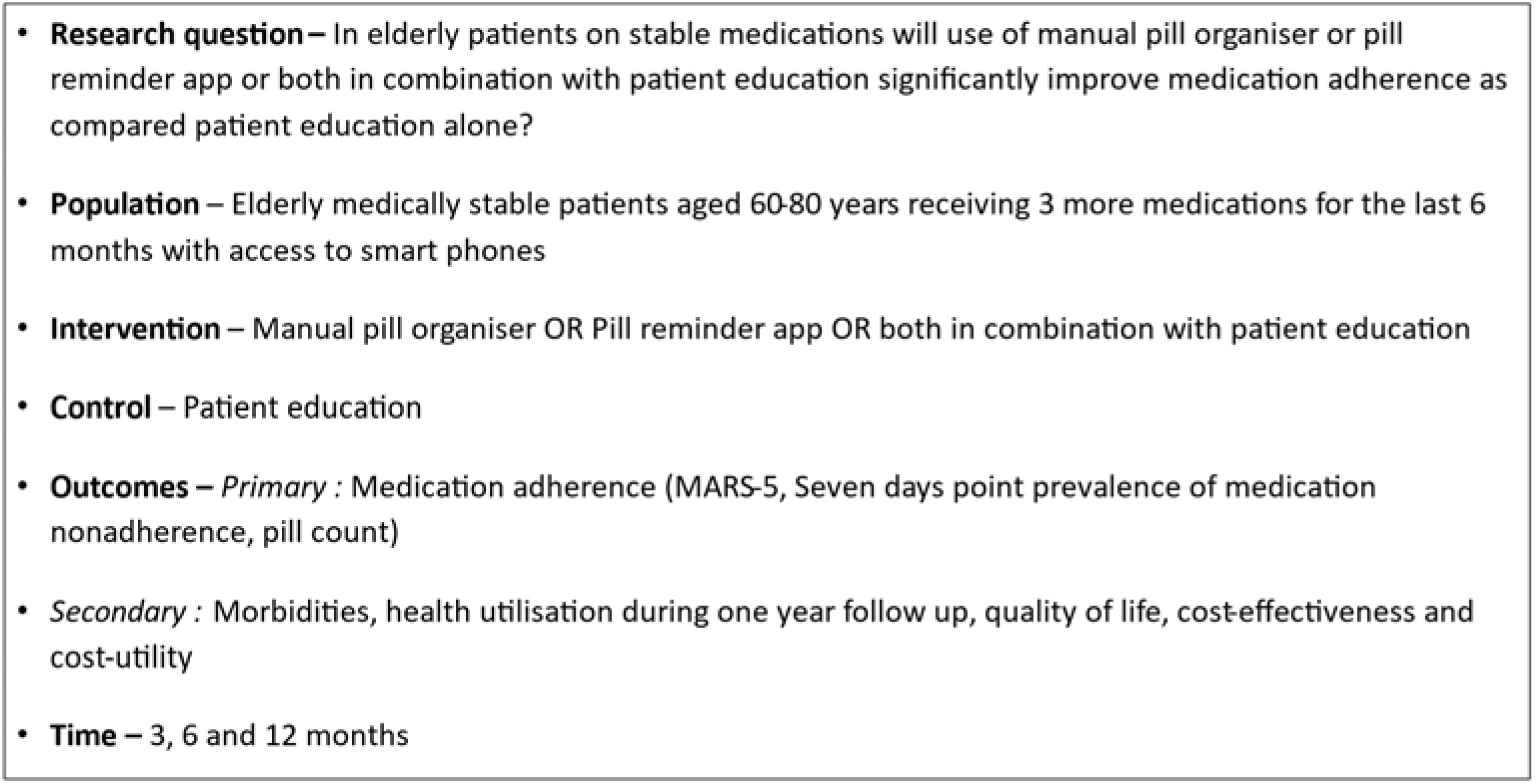
PICOT format

## Methods

### Study design

This is a 2×2 factorial design, randomised, open-label study which will be conducted across 2 different geographical locations in India. The factorial study design is depicted in Figure 2.

**Figure 2:**
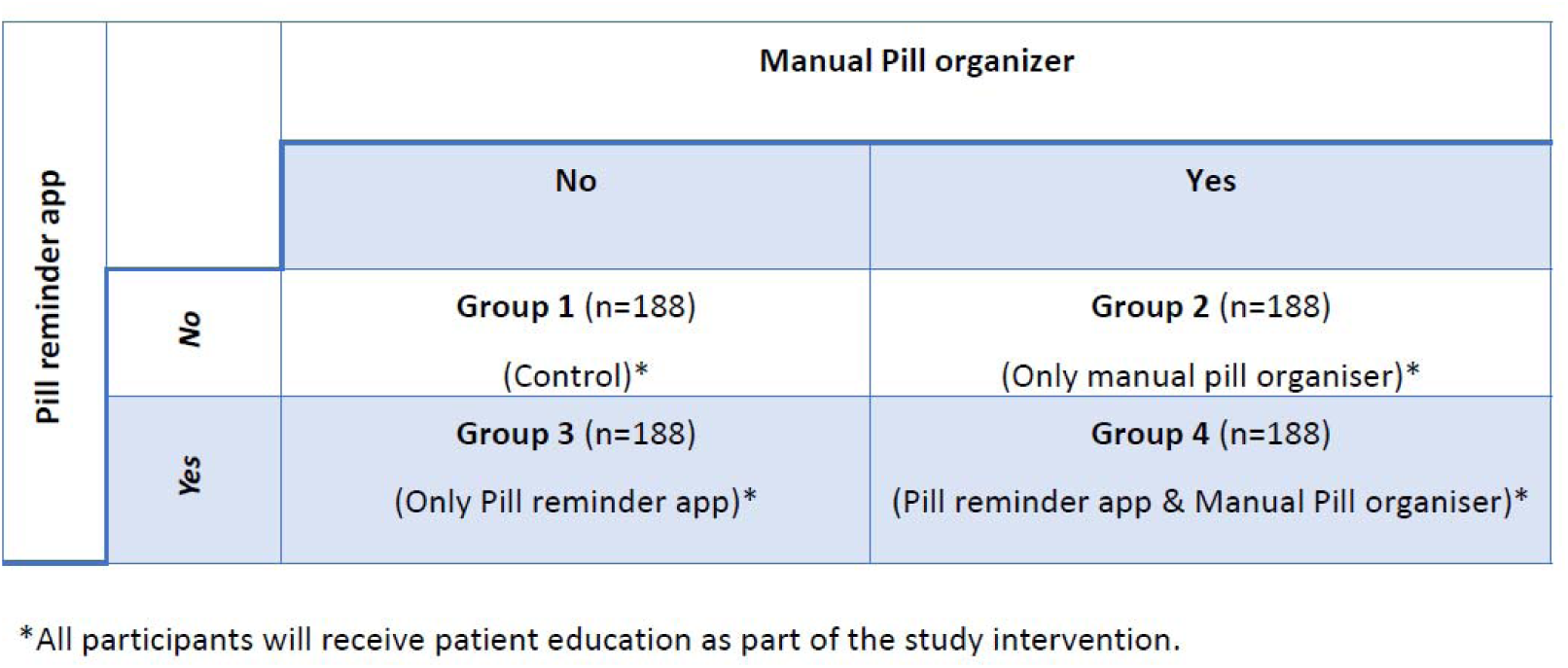
Factorial design – study interventions

### Study Settings

The study will be conducted in community-based settings amongst rural and urban populations of India. The study sites will include Vadu Rural Health Program (VRHP), KEM Hospital Research Centre, Pune (rural), Armed Forces Medical College, Pune (rural to semi-urban) and St John Medical College, Bengaluru (urban).

VRHP is a field site of KEM Hospital Research Centre, Pune, located approximately 35 km away from Pune city. It has an established health and demographic surveillance system (HDSS) that caters to around 1,75,000 population residing in 22 rural villages in the surrounding area. Armed Forces Medical College, Pune, will be another collaborating centre from Pune, and they have a service area in Kasordi village (peri-urban) near Pune city. The Department of Community Health, St John’s Medical College, Bengaluru, will conduct the study in the urban population. The department provides services through its urban health centres covering a population of 50,000, respectively, which includes a geriatric clinic and clinics for non-communicable diseases such as hypertension and diabetes.

### Study population

The study population will include elderly individuals residing within the study areas and consenting to participate in the study. Medically stable elderly individuals aged 60-80 years diagnosed with one or more non-communicable diseases and taking three or more oral medications for at least the past six months within the study area will be enumerated for enrolment. Of these, thet individuals should be able to self-administer the medications, should plan to stay in the study area for the next year, and should have access to a smartphone within the family throughout the day and the ability to use it. Individuals with a history of hospitalisation during the last month, those diagnosed with any mental or cognitive dysfunction, individuals with an expected life expectancy of less than one year (e.g. severe debility due to advanced age, advanced stage malignant cancers) or those who have used MPO or PRA for improving medication adherence in the past will be excluded. Enrolment of the eligible participants will be done by trained study staff at study clinics conducted in the vicinity of the village or through home visits. The participants will be enrolled only after written informed consent by the study physician. The informed consent documents will be translated in local languages (Marathi, Hindi and Kannada depending upon the geographical locations) and the translations will be approved by the institutional ethics committees. During the consenting process, the study procedures will be carefully explained to the study participant and their family members in in simple local language. Participants will be withdrawn from the study in case they out-migrate for more than 3 months during the study follow up or if they are not willing to be part of the study. The participants will be permitted to take all medications as per the advice of their treating physicians during the study follow up.

### Patient and Public Involvement

The study interventions have been piloted amongst patients during the initial phase of the study, and the feedback provided by the community members during interviews and focus group discussions helped to shape the study interventions, such as patient education material, and refine the manual pill organiser and pill reminder app. The protocol has been discussed with the members of the community engagement board, and their inputs in designing the community engagement sessions will be implemented. AsAs part of the dissemination plan, study findings will be shared with participants and local community members through community dissemination meetings.

### Study interventions

The participants will be randomised to one of the study groups as shown in Figure 1 and will receive one or more of the following interventions for 12 months.

### Patient Education

Patient education will involve a behavioural counselling session and education about common misbeliefs around the intake of medicines, the consequences of poor medication adherence and the importance of good medication adherence using pictorial depictions. Also, study leaflets will be handed over to the participants that explain common problems and solutions around medication non-adherence and the dos and don’ts of medication intake. The contents of the educational material have been developed through a literature search (17,18) and the findings of the pilot study. The patient education material in the form of flipcharts and patient education leaflets will be developed in English and will be translated into Marathi, Hindi, and Kannada for end users. Patient education will be provided at baseline and at the end of 6 months during the study.

### Manual pill organisers

A simple MPO that can sort medicines for up to 7 days using 3 compartments per day will be used. The box will have labels for each day and the timing of the day for ease of use. The compartments for each day would be coloured differently and can be removed from the box if needed. The MPO has been selected based on a review of available pill organisers in the Indian market while considering availability, external features, cost, ease of use, and acceptability in the pilot study. The use of pill organisers will be explained and demonstrated to the participants and their caregivers at baseline.

### Pill reminder app

An android-based PRA called “MedSathi” has been developed through an external vendor specifically for the study. The PRA has provisions for entering medicine regimes with a brand name, dose, frequency of dosing, type of formulation, and visual image of the medicine, with additional provision of maintaining a record for consumption of medicines. The PRA has an alarm and notification that can be customised depending on the medication schedule of the participant. The PRA will be installed in the smartphones of participants, and its use will be demonstrated at baseline. In case a smartphone is not available with the participant but is available with a caregiver and can be accessed throughout the day, the app will be installed on the caregiver’s smartphone. The details of the app development, challenges encountered, and feasibility of using the PRA in the elderly have been summarised elsewhere (16).

### Randomisation

Block and age-stratified randomisation will be achieved using a computer-generated randomisation list using a web-based program at the individual level, using fixed randomisation at a 1:1:1:1 ratio. The two age strata will include 60-70 years and 71-80 years. Block sizes of 4 and 8 will be used. The randomisation list will be created by a person outside the study, and the list will be included in the REDCapRedCap module for randomisation which will be accessible to the study staff responsible for allocation of enrolment. The randomisation is done centrally for the multi-centre study.

### Study outcomes

#### Medication adherence

Medication adherence will be assessed using three different methods, including the medication adherence rating system-5 [MARS-5] (19,20), self-reported seven-point prevalence of medication non-adherence, and pill count (21). MARS-5 and self-reported seven-day point prevalence of medication non-adherence will be assessed at baseline, 3, 6, and 12 months. Pill counts will be assessed towards the end of each quarter in each group by the Field Research Assistants during home visits. TheT number of pills available and the ones consumed by the participant will be calculated on two days separated by a period of 15-20 days. During this period, the participant will be informed to store the empty blister packs or containers for all medications consumed. The pill counts will be done for designated medications for non-communicable diseases and will exclude medications consumed for any ongoing acute illnesses like cold and cough. The pill count will be calculated as the number of pills taken in the given duration (the number of pills dispensed – the number of pills counted). The number of pills expected to have been taken will be calculated by multiplying the daily dose (1/2, 1, or 2 tablets) by the number of days. Successful adherence to pill counts will be defined as at least 80% of the pills taken for the given number of days (21,22). Adherence measured using the pill count method will be considered the gold standard.

#### Other outcomes

All morbidities will be recorded monthly for the participants throughout the study period. Healthcare utilisation will be measured in the form of the number of visits to health care providers (hospitalisation, emergency department visits, outpatient visits, general practitioner visits, pharmacy visits, etc) will be measured throughout the study period (23).

Health-related quality of life (HRQoL) will be assessed by the administration of a 5-level EQ-5D version (EQ-5D-5L) at baseline and 12 months. The EQ-5D-5L essentially consists of 2 parts: the EQ-5D descriptive system and the EQ visual analogue scale (EQ VAS) (24). The descriptive system comprises five dimensions: mobility, self-care, usual activities, pain/discomfort, and anxiety/depression. Each dimension has 5 levels: no problems, slight problems, moderate problems, severe problems, and extreme problems. The patient is asked to indicate his/her health state by ticking the box next to the most appropriate statement in each of the five dimensions. This decision results in a 1-digit number that expresses the level selected for that dimension. The digits for the five dimensions can be combined into a 5-digit number that describes the patient’s health state. The EQ VAS records the patient’s self-rated health on a vertical visual analogue scale, where the endpoints are labelled ’The best health you can imagine’ and ’The worst health you can imagine’. The VAS can be used as a quantitative measure of health outcomes that reflect the patient’s judgement. The summary of study outcomes is shown in Table 1.

**Table 1:** Study outcomes for the study.

| **Instrument** | **Description** |
| --- | --- |
| Medication Adherence Report Scale (MARS-5) (19,20) | 5-item self-reported scale to measure intentional and non-intentional adherence |
| Seven-day point prevalence of medication non-adherence | Number of days in the past 7 days when you missed taking the pills |
| Pill count (21) | Number of pills taken in the given duration (the number of pills dispensed – the number of pills counted). Successful adherence to pill counts will be defined as at least 80% of the pills taken during each period. |
| Morbidities | Any adverse event in the form of illness, hospitalization or adverse laboratory parameter |
| Healthcare utilization | Visits to health care providers (hospitalisation, emergency department visits, outpatient visits, general practitioner visits, pharmacy visits, etc) |
| Costing | The overall cost of developing each study intervention (i.e., patient education, MPO, and PRA) |
| Quality of life (24) | 5-level Euroqol 5D version (EQ-5D-5L) that measures mobility, self-care, usual activities, pain/discomfort, and anxiety/depression, and QOL using VAS. |
| Cost-effectiveness | The cost incurred for each intervention for unit improvement in medication adherence at the end of the study period. |
| Cost-utility | The difference in the costs per unit difference in the QALY (quality-adjusted life years). |

### Study procedures

The study flow chart is depicted in Figure 3. Elderly individuals in the study area in the eligible age group will be pre-screened at the community level for intake of multiple medications and consent for participation. Those who are willing will be invited to participate in the study. At the study visit, the participants will undergo a written informed consent. At the screening, the potential participants will be screened by a physician against the eligibility criteria, and those eligible will be enrolled in the study.

**Figure 3:**
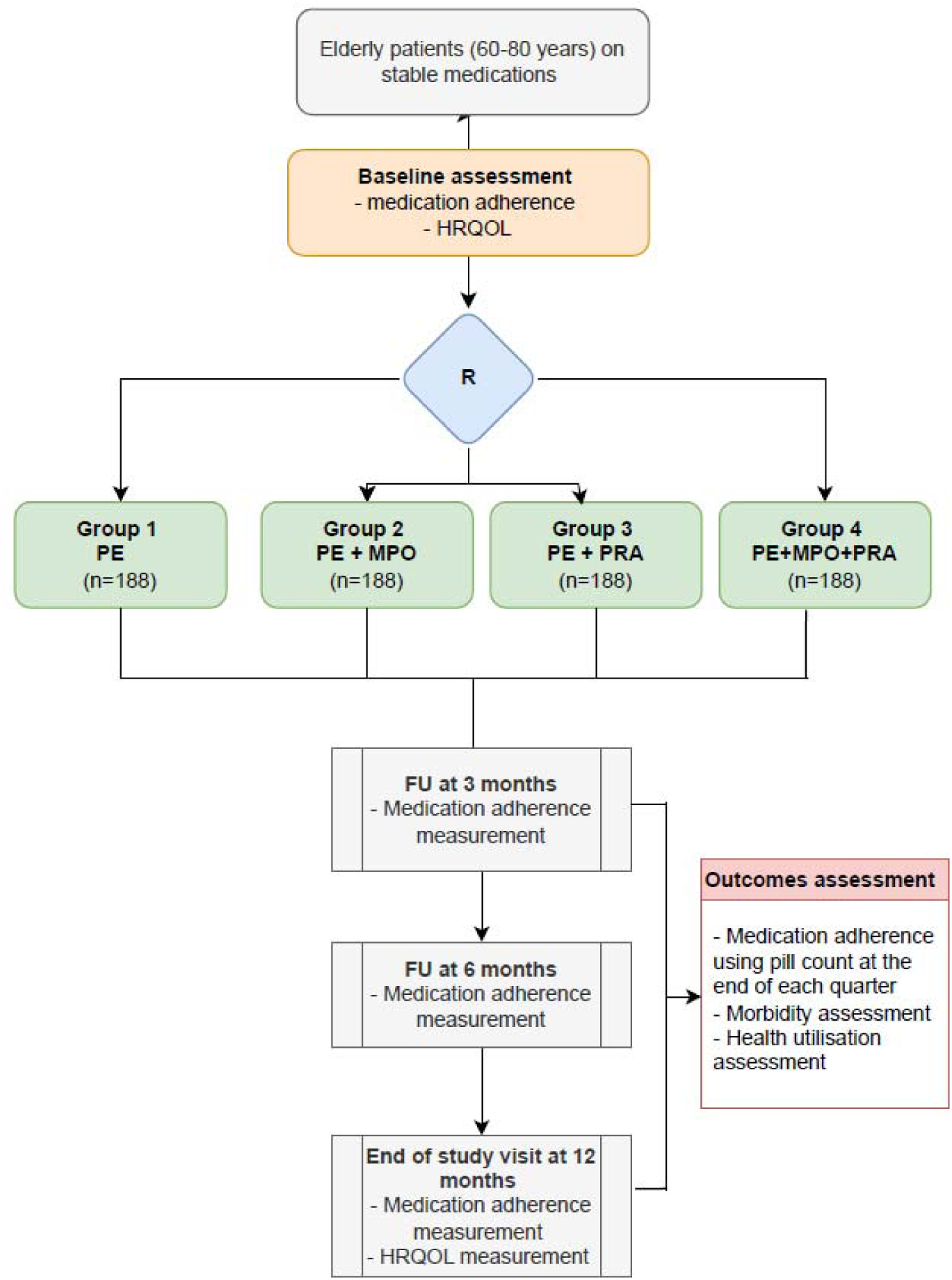
Study flow chart *PE-patient education, MPO-manual pill organiser, PRA – pill reminder app, HRQOL-health-related quality of life, FU – follow up, R-randomisation*

The participants will be followed up through monthly home visits or telephonic visits throughout the study duration for assessment of compliance with the study interventions, health, and safety outcomes. The participants will be asked to follow up with their physicians for an overall assessment of their health; however, they will be provided with medical assistance or referral if needed. There will be study visits at 3, 6, and 12 months for assessment of medication adherence, clinical outcomes, and safety assessments. Quality of life assessment will be done at baseline and the end of the study. Details of study procedures with their timelines are shown in Table 2. Community engagement activities in form of village meetings will be done through the study period to keep the local population updated about the ongoing study and to facilitate retention of participants. In case of dropouts, the data collected from the participants till the last follow up will be used for data analysis.

**Table 2:**
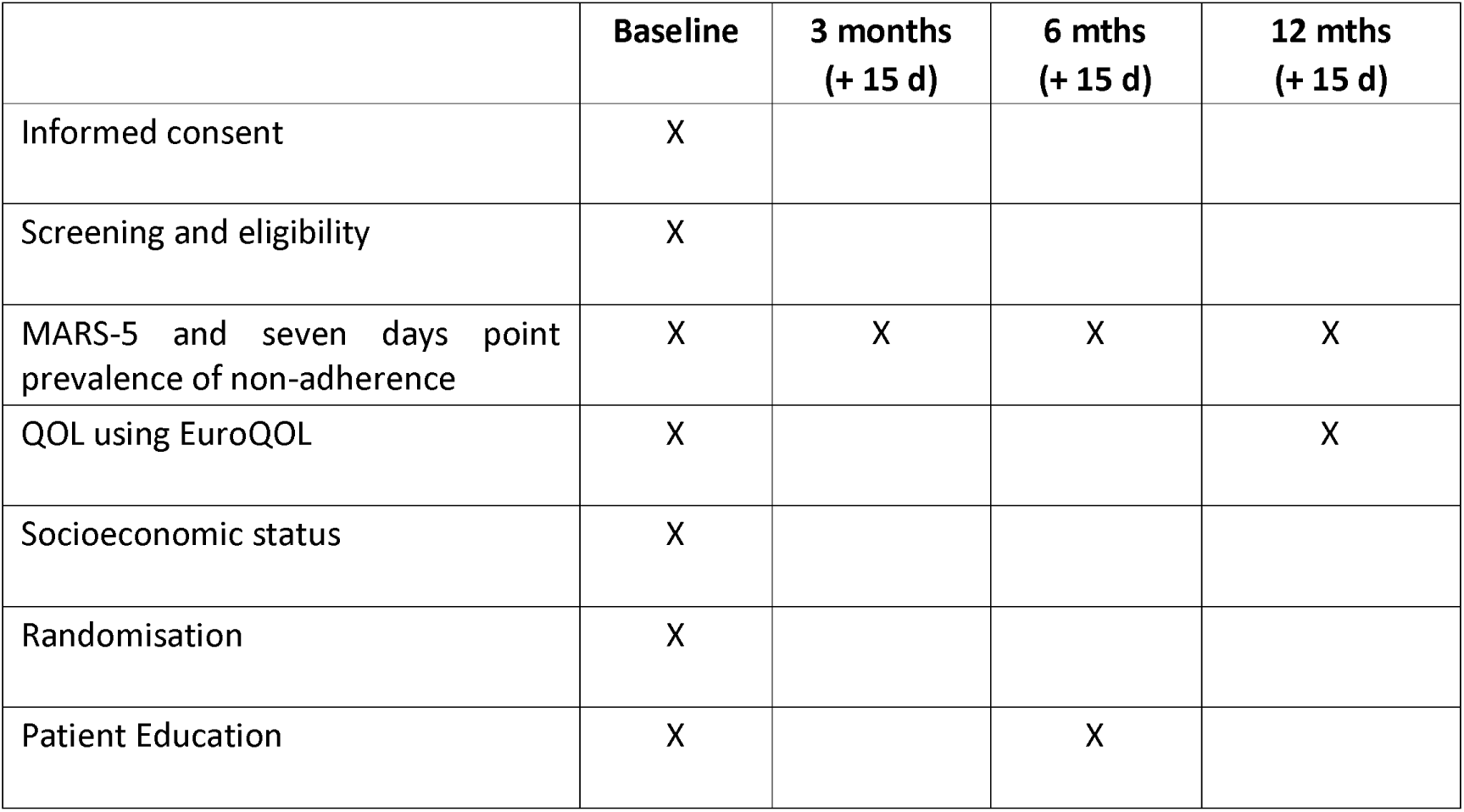
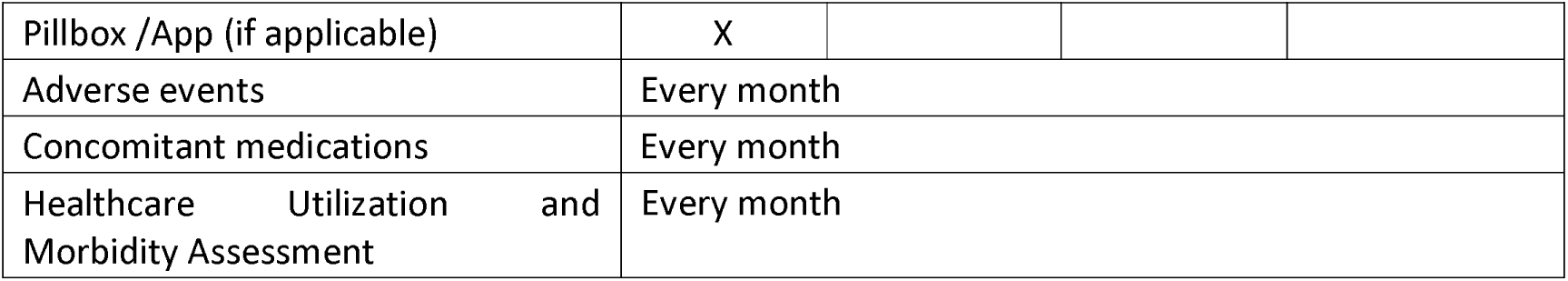
SPIRIT table for evaluation of the study.

|  | **Baseline** | **3 months**<br>**(+ 15 d)** | **6 mths**<br>**(+ 15 d)** | **12 mths**<br>**(+ 15 d)** |
| --- | --- | --- | --- | --- |
| Informed consent | X |  |  |  |
| Screening and eligibility | X |  |  |  |
| MARS-5 and seven days point prevalence of non-adherence | X | X | X | X |
| QOL using EuroQOL | X |  |  | X |
| Socioeconomic status | X |  |  |  |
| Randomisation | X |  |  |  |
| Patient Education | X |  | X |  |

Confidential
|  |  |
| --- | --- |
| Pillbox /App (if applicable) | X |
| Adverse events | Every month |
| Concomitant medications | Every month |
| Healthcare Utilization and Morbidity Assessment | Every month |

## Data management

The study data will be collected using electronic case record forms prepared in REDCap. Each site will have access to a site-specific electronic database. The identification of the study participants in the study database will be through a unique study identification number and will not contain any personal identifying information. Data quality control, storage, retrieval, security, and documentation will be ensured by the coordinating site, where the coordinating site will have access to all sites’ data. Most data quality checks will be done at the data capture. The study site teams will ensure consistency between the source data and the data present in the EDC systems. In addition, electronic edit check programs will be executed on a weekly basis to query any missing fields or through manual edits of the data by the central data management team.

After completion of data coding and resolution of all the queries in the database, the database will be declared to be complete and accurate and will be locked for final statistical analysis for primary and secondary endpoints.

## Sample size

Earlier studies from India done in adults with hypertension and diabetes have shown medication adherence of less than 50% (25,26). The available literature demonstrates significant improvement in various health outcomes, including all-cause mortality and hospitalisation, in patients with good medication adherence (70-80% or more) as compared to those with poor adherence. There is diverse evidence on the significant margin for compliance in different types of NCDs. For instance, a 10% increase in adherence to statin therapy has been linked to a 4% reduction in all-cause mortality(27). Similarly, in patients with diabetes, a 20% improvement in adherence to antidiabetic medications has been shown to result in a 0.2%-0.32% reduction in HbA1c levels, which is regarded as a clinically meaningful improvement (28). Based on this evidence, wewe aim to achieve a minimum 15% increase in medication adherence in any of the treatment groups.

Considering 50% background adherence to medication in the control group, for achieving a minimum 15% increase in adherence at 80% power and 5% alpha error, 169 individuals per group will be required. Considering the 10% dropout rate, 188 individuals will be recruited per group. The sample size will be stratified into two age strata (60-70 years and 71-80 years) with a 3:1 ratio. Considering this, a total of 752 participants will be enrolled in the study across the three sites.

## Preparation of variables for analysis

For MARS-5, responses for each question will be measured on a five-point Likert scale (1 = Always, 5 = Never), with higher scores indicating greater adherence. The total MARS-5 score will be used for the final analysis and will be represented as median with interquartile range. Medication adherence measured by pill count and seven-day point prevalence of medication non-adherence will be expressed as percentages. Morbidity profiles will be assessed as the number of hospitalisations and outpatient care visits per participant in each group.

## Calculation of health economic outcomes

The cost of the study interventions will be calculated during the planning, design, and implementation stages. The health utility score will be calculated as a preference weight based on the EQ-5D-5L and EQ-VAS and will be compared across study groups. The India-specific database for HRQoL will be used to calculate the utility index for each participant at baseline and the end of 12 months. QALY (quality-adjusted life years) will be calculated at baseline and end of the study based on the time spent in health and HRQoL. Cost-effectiveness will be calculated based on the cost incurred for each intervention for unit improvement in medication adherence at the end of the study period. The incremental cost-utility ratio will be calculated as the difference between the costs per unit difference in the QALY.

## Statistical methods for primary and secondary outcomes

Assumption of normality check will be assessed using Kolmogorov Smirnov test and Q-Q plot. The continuous variables such as age, BMI, duration of illness, etc. will be reported as mean and SD for the normally distributed continuous variables, median with interquartile range for the non-normal data across 4 study groups. The distribution of categorial variables will be reported as frequencies and percentages across 4 study groups. The baseline values of all the primary and secondary outcomes will be reported across study groups, appropriately using mean and SD for the continuous variables and frequency with percentages for the categorical variables. Statistical significance for the comparison of all the baseline characteristics including outcome parameters will be assessed using ANOVA, Kruskal Wallis test and Chi-square test appropriately.

An intention to treat analysis will be performed to evaluate the impact of MPO and PRA alone or in combination for all the primary outcome variables. The primary outcome variable of adherence rate (MARS, Self-reported medication non-adherence, pill counts) will be captured as a proportion at each time point, baseline, 3, 6 and 12 months. A mixed logistic effect model will be performed to assess the impact of MPO, PRA alone or in combination in improving medication adherence. The study groups will be considered as fixed effects, and a random effect will assess the individual-specific variability. A mixed linear or logistic effect model, as appropriate, will also be fitted for the secondary outcomes such as quality of life and hospital admission. Subsequently, covariates such as age, sex, and comorbidities will be added to the models to adjust for potential confounders. Site-specific variability will be explored and incorporated into the statistical model appropriately. The proportion of morbidity will be reported as frequency and percentage across study groups over time. The morbidity counts over time across study groups will be analyzed using mixed-effects models. The distribution of the morbidity pattern will be assessed, and by poisson or negative binomial model will be used appropriately. The proportion of health care utilisation and adverse clinical outcomes will be compared by study groups using Fisher’s exact test or chi-square test. A p-value of less than 0.05 will be considered statistically significant for all analyses. All the analyses will be conducted using Stata 18.0 version and R software, and the data analysis team will be blinded to the study group allocation.

## Study Monitoring

The study will be monitored by scientists and staff members from Indian Council of Medical Research on a period basis through visits to the study sites. The progress of the study will be evaluated through presentation of regular updates to the Technical Advisory Group for ICMR Safe and Rational Use of Medicines.

## Ethics and dissemination

The study has been registered in the Clinical Trial Registry of India (CTRI no: CTRI/2024/01/061975), and the study protocol has been approved by the respective institutional ethics committees [Approvals numbers: 2315]. The ethics approval was first obtained at KEM Hospital Research Centre, Pune, and subsequently the study was started at Vadu Rural Health Program of KEM Hospital Research Centre, Pune. The protocol version 1.1 was approved on 29 Sept 2023 by KEMHRC Ethics committee (KEMHRC/RVM/EC/1302). After protocol amendments, the final version 1.4 was approved on date 8 Oct 2024. Ethics approval has been obtained at St John Medical College, Bengaluru, and Armed Forces Medical College, Pune for the final version of the protocol. The study results will be published in an open-access peer-reviewed journal.

## Discussion

Medication non-adherence is an important public health concern among people receiving long-term medications for non-communicable diseases. The proposed study combines interventions in the form of patient education, a pill reminder app, and a pill organiser box, and assesses their impact on medication adherence. The factorial design of the study allows us to evaluate the effect of PRA and MPO alone or in combination in comparison with patient education alone in elderly patients. The android application planned to be used in the study has been locally developed into three Indian languages and has been pilot-tested in the Indian elderly population for its acceptability and feasibility.

We have used three different measures for the outcome of medication adherence: MARS-5, seven-day point prevalence of medication non-adherence, and pill count. Of these, the first two are subjective measures, whereas pill count is an objective measure that provides a precise measurement of medication adherence. While the subjective methods shed light on both intentional and non-intentional non-adherence, they are associated with subjective bias reporting. On the other hand, the objective measures, such as pill counts, are relatively precise; however, they may not inform about the causes for intentional adherence. Since we are using a multipronged approach in this study to cover both intentional and non-intentional aspects of medication non-adherence, we have used both types of measures. As per the available literature, there is no precise measure for medication adherence, and a multipronged approach is advisable for the measurement of adherence(29).

In addition to medication adherence, the study also plans to evaluate the effect on long-term morbidity and health economic outcomes through a one-year follow-up. Considering that the study would be conducted in two geographical sites across India, it is likely to provide real-world evidence on the effectiveness of the interventions for the Indian geriatric population.

Some of the limitations and challenges include limited literacy amongst elderly individuals and the unavailability of mobile phones among elderly individuals. Despite this limitation, the results of the study will have important policy implications in scaling up the use of manual pill organisers and pill reminder apps in the public health system of LMICs.

## Data Availability

The anonymised participant dataset, study protocol and statistical plan of analysis will be made available to interested researchers after analysis of primary study results and with prior authorisation from the study investigators.

## Status and collaboration

This study is planned as a part of the Indian Council of Medical Research (ICMR)– Safe and Rational Use of Medicines Taskforce (SRUM) and is funded by ICMR, New Delhi, India. The study is being coordinated by KEM Hospital Research Centre, Pune, India. The study was initiated Nov 2024.

## Acknowledgement

We would like to acknowledge the contributions of the members of the Central Coordinating Unit team and the Technical Advisory Group of the Indian Council of Medical Research - National Task Force for Safe and Rational Use of Medicines (SRUM NTF). Their support and assistance in project coordination, technical advice, and administrative support are greatly appreciated. We acknowledge support from the drcsystems team for their support in developing the Android-based mobile app MedSathi for the study.

## Author Contribution

AA, DA, JJC, and SR decided to publish the paper. AA has drafted the study protocol, and FNF, DA, JJS, and SR provided input to the trial design. AA, DA, RP, FNF, RB, RN, BS, PS, and HS helped plan the trial and are responsible for the acquisition of data. AA drafted the manuscript. AA, FNF, and BS lead the clinical teams in their respective institutes. SS and AA contribute to statistical analysis. All authors critically revised the manuscript. All authors are contributing to the conduct of the trial.

## References

1. Menon GR, Yadav J, John D. Burden of non-communicable diseases and its associated economic costs in India. Social Sciences and Humanities Open. 2022 Jan 1;5(1). Available from: 10.1016/j.ssaho.2022.100256

2. Kleinsinger F. The Unmet Challenge of Medication Nonadherence. Perm J. 2018;22:18–033.

3. Doggrell SA. Adherence to medicines in the older-aged with chronic conditions: does intervention by an allied health professional help? Drugs Aging. 2010 Mar 1;27(3):239–54.

4. Kengne AP, Brière JB, Zhu L, Li J, Bhatia MK, Atanasov P, et al. Impact of poor medication adherence on clinical outcomes and health resource utilization in patients with hypertension and/or dyslipidemia: systematic review. Expert Rev Pharmacoecon Outcomes Res [Internet]. 2024 Jan 2;24(1):143–54. Available from: 10.1080/14737167.2023.2266135

5. Walsh CA, Cahir C, Tecklenborg S, Byrne C, Culbertson MA, Bennett KE. The association between medication non-adherence and adverse health outcomes in ageing populations: A systematic review and meta-analysis. Vol. 85, British Journal of Clinical Pharmacology. Blackwell Publishing Ltd; 2019. p. 2464–78.

6. Khan R, Socha-Dietrich K. Investing in medication adherence improves health outcomes and health system efficiency: Adherence to medicines for diabetes, hypertension, and hyperlipidaemia. OECD Health Working Papers [Internet]. 2018;(105). Available from: 10.1787/8178962c-en

7. Junaid Farrukh M, Makmor Bakry M, Hatah E, Hui Jan T. Medication adherence status among patients with neurological conditions and its association with quality of life. Saudi Pharmaceutical Journal. 2021 May 1;29(5):427–33.

8. Peacock E, Joyce C, Craig LS, Lenane Z, Holt EW, Muntner P, et al. Lowmedication adherence is associatedwith decline in health-related quality of life: Results of a longitudinal analysis among older women andmen with hypertension. J Hypertens. 2021 Jan 1;39(1):153–61.

9. Bhattacharya D, Aldus CF, Barton G, Bond CM, Boonyaprapa S, Charles IS, et al. The feasibility of determining the effectiveness and cost-effectiveness of medication organisation devices compared with usual care for older people in a community setting: Systematic review, stakeholder focus groups and feasibility randomised controlled . Health Technol Assess (Rockv). 2016;20(50):1–127.

10. Doggrell SA. Adherence to medicines in the older-aged with chronic conditions: Does intervention by an allied health professional help? Drugs Aging. 2010;27(3):239–54.

11. Bae SG, Kam S, Park KS, Kim KY, Hong NS, Kim KS, et al. Factors related to intentional and unintentional medication nonadherence in elderly patients with hypertension in rural community. Patient Prefer Adherence. 2016 Sep 29;10:1979–89.

12. Kini V, Michael Ho P. Interventions to Improve Medication Adherence: A Review. JAMA - Journal of the American Medical Association. 2018;320(23):2461–73.

13. Doggrell SA. Adherence to Medicines in the Older-Aged with Chronic Conditions Does Intervention by an Allied Health Professional Help? Available from: doi: 10.2165/11532870-000000000-00000.

14. Conn VS, Ruppar TM, Chan KC, Dunbar-Jacob J, Pepper GA, De Geest S. Packaging interventions to increase medication adherence: Systematic review and meta-Analysis. Curr Med Res Opin. 2015;31(1):145–60.

15. Armitage LC, Kassavou A, Sutton S. Do mobile device apps designed to support medication adherence demonstrate efficacy? A systematic review of randomised controlled trials, with meta-analysis. BMJ Open. 2020 Jan;10(1):e032045. 10.1136/bmjopen-2019-032045

16. Nimkar R BPRADAA. Evaluation of acceptability and feasibility of using manual pill organizers and pill reminder apps for improving medication adherence amongst elderly population from rural Maharashtra. Indian Journal of Pharmacology (Inpress). 2024;

17. Hugtenburg JG, Timmers L, Elders PJ, Vervloet M, van Dijk L. Definitions, variants, and causes of nonadherence with medication: a challenge for tailored interventions. Patient Prefer Adherence. 2013;7:675–82.

18. Medication Adherence: We Didn’t Ask and They Didn’t Tell. Fam Pract Manag. 2013;20(2):25–30.

19. Molloy GJ, Messerli-Bürgy N, Hutton G, Wikman A, Perkins-Porras L, Steptoe A. Intentional and unintentional non-adherence to medications following an acute coronary syndrome: A longitudinal study. J Psychosom Res. 2014;76(5):430–2.

20. Chan AHY, Horne R, Hankins M, Chisari C. The Medication Adherence Report Scale: A measurement tool for eliciting patients’ reports of nonadherence. Br J Clin Pharmacol. 2020;86(7):1281–8.

21. Lee JK, Grace KA, Foster TG, Crawley MJ, Erowele GI, Sun HJ, et al. How should we measure medication adherence in clinical trials and practice? Ther Clin Risk Manag. 2007;3(4):685–90.

22. Juste M. Medication adherence. Pharmacien Hospitalier et Clinicien. 2019;54(2):107–8.

23. Walsh CA, Cahir C, Tecklenborg S, Byrne C, Culbertson MA, Bennett KE. The association between medication non-adherence and adverse health outcomes in ageing populations: A systematic review and meta-analysis. Br J Clin Pharmacol. 2019;85(11):2464–78.

24. Group E, Eq- T, Vas EQ. EQ-5D-5L | About. 2009;23–5.

25. Mishra R, Sharma SK, Verma R, Kangra P, Dahiya P, Kumari P, et al. Medication adherence and quality of life among type-2 diabetes mellitus patients in India. World J Diabetes. 2021;12(10):1740–9.

26. Medi R, Mateti U, Kanduri K, Konda S. Medication adherence and determinants of non-adherence among south Indian diabetes patients. Journal of Social Health and Diabetes. 2015;03(01):048–51.

27. Chodick G, Rotem RS, Miano TA, Bilker WB, Hennessy S. Adherence with statins and all-cause mortality in days with high temperature. Pharmacoepidemiol Drug Saf. 2024 Jun 1;33(6). doi: 10.1002/pds.5817.

28. García-Pérez LE, Álvarez M, Dilla T, Gil-Guillén V, Orozco-Beltrán D. Adherence to therapies in patients with type 2 diabetes. Vol. 4, Diabetes Therapy. Springer Healthcare; 2013. p. 175–94.

29. Lam WY, Fresco P. Medication Adherence Measures: An Overview. Vol. 2015, BioMed Research International. Hindawi Publishing Corporation; 2015. doi: 10.1155/2015/217047.

